# Brain disconnectome mapping and serum neurofilament light levels in multiple sclerosis

**DOI:** 10.1101/2021.04.21.21255887

**Authors:** Henning H. Rise, Synne Brune, Claudia Chien, Tone Berge, Steffan D. Bos, Magi Andorra, Irene Pulido Valdeolivas, Mona K. Beyer, Piotr Sowa, Michael Scheel, Alexander U. Brandt, Susanna Asseyer, Kaj Blennow, Mads L. Pedersen, Henrik Zetterberg, Michel Thiebaut de Schotten, Maria Cellerino, Antonio Uccelli, Friedemann Paul, Pablo Villoslada, Hanne F. Harbo, Lars T. Westlye, Einar A. Høgestøl

## Abstract

The pathophysiological mechanisms for classical plaque characteristics and their predictive value for clinical course and outcome in multiple sclerosis is unclear. Connectivity-based approaches incorporating the distribution and magnitude of the extended brain network aberrations caused by lesions may offer higher sensitivity for axonal damage. Using individual brain disconnectome mapping, we tested the longitudinal associations between putative brain network aberrations and levels of serum neurofilament light chain (sNfL) as a neuroaxonal injury biomarker.

Multiple sclerosis patients (*n* = 328, mean age 42.9 years, 71 % female) were prospectively enrolled at four European multiple sclerosis centres, and reassessed after two years (n = 280). Post-processing of 3 Tesla (3T) MRI data was performed at one centre using a harmonized pipeline, and disconnectome maps were calculated using BCBtoolkit based on individual lesion maps. Global disconnectivity (GD) was defined as the average disconnectome probability in each patient’s white matter. Serum NfL concentrations were measured by single molecule array (Simoa). Robust linear mixed models (rLMM) with GD or T2-lesion volume (T2LV) as dependent variables, patient and centre as a random factor, sNfL, age, sex, timepoint for visit, diagnosis, and treatment as fixed factors were run.

Robust LMM revealed significant associations between higher levels of GD and increased sNfL (*t* = 2.30, *β* = 0.03, *p* = 0.02), age (*t* = 5.01, *β* = 0.32, *p* < 5.5 × 10^−7^), and diagnosis progressive multiple sclerosis (PMS); *t* = 1.97, *β* = 1.06, *p* = 0.05), but not for sex (*t* = 0.78, *p* = 0.43), treatments (effective; *t* = 0.85, *p* = 0.39, highly-effective; *t* = 0.86, *p* = 0.39) or sNfL change between base line and two-year follow up (*t* = −1.65, *p* = 0.10). Voxel-wise analyses revealed distributed associations in cerebellar and brainstem regions.

In our prospective multi-site multiple sclerosis cohort, rLMMs demonstrated that the extent of global brain disconnectivity is sensitive to a systemic biomarker of axonal damage, sNfL, in patients with multiple sclerosis. These findings provide a neuropathological correlate of advanced disconnectome mapping and provide a platform for further investigations of the functional and clinical relevance in patients with brain disorders.

## Introduction

MRI of the brain and spinal cord is essential for diagnostics and clinical management of patients with multiple sclerosis, a chronic autoimmune disorder of the central nervous system.^1, 2^ MRI technology improvements have enabled highly accurate visualization of white matter lesions due to multiple sclerosis-related pathological processes.^3, 4^ While T2-weighted lesions represent validated MRI markers in multiple sclerosis, their neuroanatomical and pathophysiological implications are complex, and their predictive value for clinical trajectories and outcomes has been modest. ^3-5^ This clinico-radiological-paradox may be alleviated by imaging methods sensitive to subtle primary or secondary axonal damage extending beyond the visual foci of the lesions.^6, 7^

Recent advances in brain imaging post-processing techniques probing the human brain connectome have allowed for in vivo investigations of the distributed network-level aberrations caused by focal lesions.^8, 9^ By calculating the probability that white matter fibres, as identified in healthy individuals intersect lesions observed in the brain of multiple sclerosis patients, individual disconnectome maps offer an opportunity to conceptualize and characterize the extent of brain network aberrations due to local T2-hyperintense lesions. However, the neurobiological and pathological correlates of disconnectome mapping remain unknown, partly due to the non-invasive nature of the procedures and the lack of accurate biomarkers reflecting axonal damage.

Over the last two decades, central (CSF) and peripheral (plasma and serum) neurofilaments have gained increased attention as candidate biomarkers of neuroaxonal injury as these structural scaffolding proteins are exclusively expressed in neurons and released into the periphery upon axonal damage with potential to monitor subclinical disease activity in neurodegenerative disorders such as multiple sclerosis. Neurofilament light chain (NfL) is a subunit of neurofilaments, and its concentration increase both of neurofilament protein in CSF and blood proportionally to the degree of axonal damage.^10^ Recent studies employing the single molecule array (Simoa®) technique have demonstrated that NfL concentrations in serum and CSF are highly correlated, enabling the use of serum NfL (sNfL) as a reliable biomarker reflecting axonal injury.^11, 12^ Higher levels of serum or CSF NfL have been associated with MRI lesions, spinal cord and brain atrophy, gadolinium-enhancing lesions, increasing age, recent clinical relapses, clinical disability, and treatment efficacy in multiple sclerosis.^12-22^ Recent studies based on national health registry studies proposed NfL as a potential marker for predicting multiple sclerosis risk and disease course, even at the earlier stages of the disease in subjects with radiologically isolated syndrome (RIS) and clinically isolated syndrome (CIS).^21, 23^ Combining NfL with advanced neuroimaging measures may enable increased understanding of multiple sclerosis pathophysiology and improved prognosis prediction, and better, as well as treatment responses monitoring.^21, 22, 24-26^

In this longitudinal multicenter study comprising a large real-world heterogeneous multiple sclerosis cohort (*n* = 328), we tested for associations between state-of-the-art brain disconnectivity mapping and sNfL concentrations. To leverage the combined cross-sectional and longitudinal study design, we used robust linear mixed-effects models (rLMM) including relevant covariates to test for overall associations between global disconnectivity and sNfL concentrations as well as the interactions between sNfL concentration and changes in sNfL on global disconnectivity. Subsequently, we performed voxel-wise analyses to map the neuroanatomical distribution of associations and to compare global disconnectivity with conventional measures, employing similar rLMM testing for associations with conventional T2 lesion volume (T2LV).

## Materials and methods

### Study population

A total of 328 multiple sclerosis patients were prospectively enrolled at four European multiple sclerosis centres from July 2016 to December 2017 (68 subjects from Hospital Clinic of Barcelona, Spain; 95 subjects from Oslo University Hospital, Norway; 73 subjects from Charité-Universitaetsmedizin Berlin, Germany; 92 subjects from Ospedale Policlinico San Martino, Genoa, Italy). All multiple sclerosis patients were invited for a two-year follow-up between January 2017 and March 2020, resulting in 280 subjects (85 %) completing the longitudinal study (Table 1). All patients provided demographic information, personal multiple sclerosis history, blood samples, and assessment of expanded disability status scale (EDSS).

**Table 1.** Demographic, clinical and biomarker information of the multiple sclerosis cohort.

|  | Baseline | Follow-up |
| --- | --- | --- |
| **(A) Demographic characteristics** | *n* = 312 | *n* = 242 |
| Female % | 70.5 | 69.4 |
| Age, mean years (*SD*, *range*) | 42.9 (9.9, 19-68) | 45.1 (9.8, 21-70) |
| Disease duration, mean years (*SD*, *range*) | 11.0 (8.2, 0-43) | 13.5 (8.5, 2-46) |
| Follow-up time, mean years (*SD*, *range*) |  | 1.97 (0.34, 0.7-3.3) |
| Age at first symptom, mean years (*SD*, *range*) | 31.4 (8.9, 7-56) |  |
| *Centre (included patients)* |  |  |
| Barcelona % (*n*) | 18 (59) | 18 (44) |
| Oslo % (*n*) | 30 (95) | 33 (78) |
| Berlin % (*n*) | 22 (70) | 18 (44) |
| Genova % (*n*) | 28 (88) | 31 (76) |
| **(B) Disease modifying treatment** | *n* = 311 | *n* = 235 |
| No treatment % (*n*) | 30 (94) | 26 (61) |
| Effective treatment % (*n*) | 44 (138) | 37 (87) |
| Highly-effective treatment % (*n*) | 26 (79) | 37 (87) |
| **(C) Clinical evaluation and biomarkers** |  |  |
| *Multiple sclerosis classification* |  |  |
| CIS % (*n*) | 1.6 (5) | 1.7 (4) |
| RRMS % (*n*) | 81.4 (254) | 78.5 (190) |
| SPMS % (*n*) | 8.0 (25) | 10.3 (25) |
| PPMS % (*n*) | 9.0 (28) | 9.5 (23) |
| *Neurological disability* |  | *n* = 275 |
| EDSS, median (*IQR*, *range*) | 2.0 (2.0, 0-8) | 2.0 (2.5, 0-8) |
| Δ EDSS improving ($\leq 1$), % (*n*) | | 19 (51) |
| Δ EDSS stable ($\Delta$ EDSS $>1$), % (*n*) | | 65 (181) |
| Δ EDSS worsening ($\geq 1$), % (*n*) | | 16 (43) |
| T2 lesion volume ml, mean (*SD*, *range*) | 8.5 (10.9, 0.1-64.5) | 9.6 (11.1, 0.1-57) |
| Normalized brain volume ml, mean (*SD*, *range*) | 1507 (90, 1263-1724) | 1453 (70, 1244-1666) |
| Serum neurofilament light levels, mean pg/ml (*SD*, *range*) | 9.0 (7.2, 2.2-93.2) | 8.7 (5.5, 2.3-45.5) |
| *CIS = clinically isolated syndrome; EDSS = expanded disability status scale; IQR = interquartile range; PPMS = primary-progressive multiple sclerosis; RRMS = relapsing-remitting multiple sclerosis; SD = standard deviation; SPMS = secondary-progressive multiple sclerosis* |  |  |

Inclusion criteria included age 18-80 years, CIS or multiple sclerosis diagnosis according to the 2010 McDonald’s criteria,^27^ disease duration ≤ 15 years. Exclusion criteria were use of corticosteroids the last 30 days or a relapse in the month prior to inclusion, subjects not eligible for a blood draw, chronic diseases other than multiple sclerosis, and pregnancy during the course of the study. For patients previously treated with disease modifying treatments (DMT), a washout of at least three months was required (six months for ocrelizumab/rituximab; one year following alemtuzumab). Patients on DMTs were also included and needed to be stable for at least one year when treated with interferon (IFN)-beta or glatiramer acetate (GA) or at least six months for other treatments. 59 age-and sex-matched healthy controls (HC) (Supplementary Table 1) were recruited from the same four European multiple sclerosis centres.

### Standard protocol approvals, registrations, patient consents

The Sys4MS project was approved by the IRBs of University of Genoa, University of Oslo (REC ID: 2011/1846 A), Charité-Universitaetsmedizin, and Hospital Clinic of Barcelona. Patients provided signed informed consent prior to their enrolment on the study according to the Declaration of Helsinki.

### Serum Neurofilament light analysis

Serum samples were collected with 4 mL Vacuette Z Serum Clot Activator Tube® (Greiner bio-one International) and processed within one hour by centrifugation at 2000g for 10 minutes at 4°C. Serum aliquots were immediately stored at −80°C until analysis. Samples were thawed only once during the processing. Measurement of sNfL samples was performed in the Clinical Neurochemistry Laboratory at the Sahlgrenska University Hospital, Sweden, by board certified laboratory technicians blind to clinical data, using an ultrasensitive Single molecule array (Simoa) assay described elsewhere.^12^ A single batch of reagents was used; intra-assay coefficients of variation were below 10 % for all analyses. Two QC samples were run in duplicates in the beginning and the end of each run, repeatability and intermediate precision were 9.0 % at 25 pg/mL and 5.2 % at 80 pg/mL.

### MRI acquisition

Images were acquired at all European multiple sclerosis centres. From Centre 1 (Barcelona), a 3D magnetization prepared rapid gradient echo (MPRAGE) sequence, including the upper cervical cord (0.86 x 0.86 x 0.86 mm resolution, *repetition time (TR)* = 1970 ms, *echo time (TE)* = 2.41 ms), an axial T1-weighted post-gadolinium contrast agent sequence (0.31 x 0.31 x 3 mm resolution, *TR* = 390 ms, *TE* = 2.65 ms), and a 3D fluid-attenuated inversion recovery (FLAIR) sequence, including the upper cervical cord (1 x 1 x 1 mm resolution, *TR* = 5000 ms, *TE* = 393 ms) were acquired longitudinally using a Tim Trio MRI (Siemens Medical Systems, Erlangen, Germany), and MAGNETOM Prisma MRI (Siemens Medical Systems) at the follow-up assessment from January 15^th^ 2018. From Centre 2 (Oslo), a 3D sagittal brain volume (BRAVO) sequence for pre- and post-gadolinium contrast agent administration, including the upper cervical cord (1 x 1 x 1 mm resolution, *TR* = 8.16 ms, *TE* = 3.18 ms), and a 3D FLAIR sequence, including the upper cervical cord (1 x 1 x 1.2 mm resolution, *TR* = 8000 ms, *TE* = 127.25 ms) were acquired longitudinally using a Discovery MR750 MRI (GE Medical Systems). From Centre 3 (Berlin), a 3D sagittal MPRAGE sequence, including the upper cervical cord (1 x 1 x 1 mm resolution, *TR* = 1900 ms, *TE* = 3.03 ms), and a 3D FLAIR sequence, including the upper cervical cord (1 x 1 x 1 mm resolution, *TR* = 6000 ms, *TE* = 388 ms) were acquired longitudinally using a Tim Trio MRI (Siemens Medical Systems, Erlangen, Germany). From Centre 4 (Genova), a sagittal fast-spoiled gradient-echo (FSPGR) sequence, including the upper cervical cord (1 x 1 x 1 mm resolution, *TR* = 7.31 ms, *TE* = 3.00 ms), a 3D turbo field echo (TFE) sequence for post-gadolinium contrast agent administration (1 x 1 x 1 mm resolution, *TR* = 8.67 ms, *TE* = 4.00 ms), and a 3D FLAIR sequence, including the upper cervical cord (1 x 1 x 1 mm resolution, *TR* = 6000 ms, *TE* = 122.16 ms) were acquired longitudinally using a Signa HDxt MRI (GE Medical Systems) and Ingenia MRI (Philips Medical Systems) at baseline and MAGNETOM Prisma MRI (Siemens Medical Systems) for follow-up assessment.

### MRI pre- and post-processing at Berlin reading centre

Pre-processing included alignment to Montreal Neurological Institute (MNI) −152 standard space (using fslreorient2std), white and grey matter brain masking (using Computational Anatomy Toolbox 12 Toolbox for MATLAB),^28^ N4-bias field correction (Advanced Normalization Tools, http://stnava.github.io/ANTs/) and linear, rigid body registration of T2-weighted (FLAIR) images to T1-weighted (MPRAGE, BRAVO, and FSPGR) images (FSL FLIRT).^29, 30^ T1-weighted and FLAIR follow-up scans were co-registered to the individual first session using the transformation matrices saved from the first session transformation from native space images to MNI-152 standard space using FSL FLIRT. Post-contrast agent T1-weighted images were also co-registered to MNI-152 standard space and longitudinally when available.

T2-hyperintense lesion segmentation was performed using a semi-automated pipeline at one centre on co-registered T1-weighted images and T2-weighted FLAIR images by two experienced MRI technicians. Lesions were segmented and saved as binary masks using ITK-SNAP (www.itksnap.org).^31^ First session lesion masks were subsequently overlayed onto the second session co-registered T1-weighted and FLAIR images for editing, to include any T2-hyperintense lesion changes (i.e., new lesions, enlarging lesions, or decreasing lesions) in the follow-up scans. Any discrepancies in co-registrations that were visible between sessions were corrected manually using the ITK-SNAP automated registration tool before follow-up lesion mask edits. Binary gadolinium enhancing lesion masks were created manually using the same tools on the post-gadolinium T1-weighted MR images by the same two technicians. Lesion counts and volumes were extracted from lesion masks using fslmaths (https://fsl.fmrib.ox.ac.uk/fsl/fslwiki/Fslutils).T2-hyperintense lesion masks were used to fill longitudinally co-registered T1-weighted (not post-gadolinium scans) images using the FSL lesion filling tool (https://fsl.fmrib.ox.ac.uk/fsl/fslwiki/lesion_filling), utilizing white matter masks created from the Computational Anatomy Toolbox for SPM12 (CAT12, http://www.neuro.uni-jena.de/cat/). Lesion-filled T1-weighted images were then used for whole-brain white and grey matter volume extraction, including the follow-up session percent brain tissue volume change (PBVC), normalised for subject head size, was estimated with SIENAX,^32^ part of FSL.^33^

### Disconnectome maps

Disconnectome maps were calculated using BCBtoolkit.^34^ This approach uses diffusion-weighted imaging data from 10 healthy controls^35^ derived from the 7T tractography made available in de Schotten et al., Nat Commun, 2020^8^ to track fibers passing through each lesion.^36^ Patients’ lesions in the MNI152 space were registered to each control native space using affine and diffeomorphic deformations^37, 38^ and subsequently used as seed for the tractography in Trackvis.^39^ The resulting tractograms were transformed to visitation maps,^36^ binarised, and brought to MNI152 space using the inverse of model deformations. Finally, a percentage overlap map was computed by summing at each voxel in MNI space the normalized visitation map of each healthy subject. A white matter mask was made by including all non-zero values across the individual disconnectome maps and subsequently used in analysis of voxel-wise and global effects of disconnectome maps.

### Global and regional disconnectome

Global disconnectome was calculated for each patient by computing the average disconnectome score across all white matter voxels. Voxel-wise analysis was performed using FSL randomise^40^ on disconnectome maps from all patients at inclusion. The general linear model (GLM) applied to randomise was designed with a single-group average disconnectome map as response variable with sNfL as explanatory variable, and with age, sex, diagnosis, and treatment as additional covariates in the main design matrix. Permutation-based inference was performed across 5000 iterations for both contrasts (positive and negative associations with sNfL) with threshold-free cluster enhancement (TFCE).^41^

### Statistical analysis

Analyses were performed using R 4.0.3.^42^ To investigate longitudinal associations between sNfL and GD and T2LV from inclusion to two-year follow-up, two separate rLMM were conducted examining GD or T2LV as dependent variables, using rlmer from the robustlmm R package.^43^ All continuous variables were standardized before running the analyses. All models included age, sex, diagnosis, timepoint of visit and treatment as fixed effect terms, and subject identifier (ID) as random effects.

### Data availability

Anonymized data is available through the MultipleMS EU project and database (www.multiplems.eu) upon registration.

## Results

### Participant demographics and characteristics

At baseline, the multiple sclerosis cohort consisted of 71 % women, 83 % CIS and relapsing and remitting-remitting multiple sclerosis (RRMS) patients, with a mean age of 42.9 years (Table 1). The mean disease duration was 11 years and 30 % of the multiple sclerosis patients were untreated, while 44 % and 26 % were using effective and highly-effective DMTs, respectively. The multiple sclerosis subjects who completed follow-up were re-examined on average 2.0 years after baseline assessment (*range* = 0.7-3.3 years). At follow-up, 37 % of the multiple sclerosis subjects were using effective treatment, and 37 % highly-effective DMTs, while the proportion of multiple sclerosis subjects currently not using DMTs was decreased to 26 %. For both time points, median EDSS was 2.0 (*Interquartile range (IQR)* = 2.0 at baseline and *IQR* = 2.5 at follow-up, *range* = 0-8 years). Serum NfL levels at baseline were on average 8.9 pg/ml (*SD* ± 4.4 pg/ml) for those with multiple sclerosis and 7.0 pg/ml (*SD* ± 3.8) for HCs. The correlation between GD and T2LV was very high (*r* = 0.80) (Supplementary Fig. 1). For our study, 312 multiple sclerosis patients (95 %) met all criteria with available clinical, MRI, and sNfL data at baseline, while 242 multiple sclerosis patients (86 %) fulfilled the requirements at follow-up.

### Lesion and disconnectome maps

Figure 1 shows a probabilistic map representing the overlap of disconnectome across all included subjects in addition to selected individual maps. In the disconnectome map, the value in each voxel takes into account the interindividual variability of tract reconstructions in controls, and indicate a probability of disconnection from 0 to 100 % for a given lesion.^9^ We thresholded the resulting map at >50 %, indicating that at least half of the individuals in the training set have trajectories that intersect the lesion location in each patient for the corresponding fibers to be part of the patients’ disconnectome maps.

**Figure 1.**
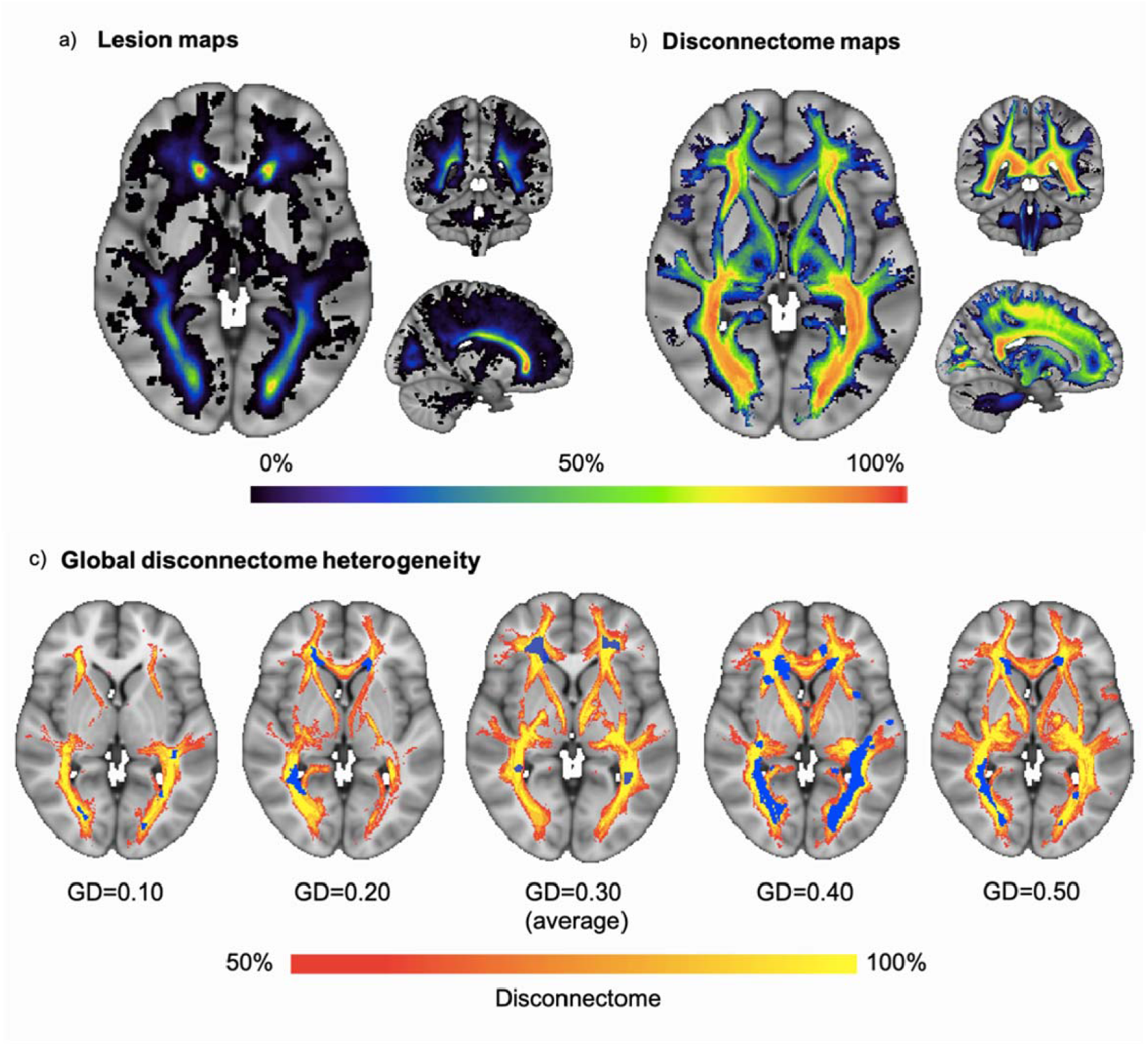
Probability distribution heterogeneity of lesions and disconnectome in the multiple sclerosis sample. An overview with representative example slices of the probability distribution of **(A)** lesion and **(B)** disconnectome maps, as seen in a large axial slice to the left and smaller coronal and sagittal slices on the top right and lower right, respectively. Masked out in both **(A)** and **(B)** are the borders of the resulting global masks and the colour-filled probability maps corresponding to the percentage of subjects with a lesion or disconnectome maps in the depict voxels. The corresponding colour bar is shown at the bottom of the figure. In **(C)** we depicted five subjects with different levels of GD, with blue colour highlighting the underlying lesion masks, while the red to yellow colour indicates the probability of disconnectivity.

### Longitudinal analyses with robust linear mixed models

Table 2 summarizes the results from the rLMM testing for associations between GD or T2LV with sNfL, timepoint, age, sex, treatment and multiple sclerosis phenotype as covariates. Briefly, the GD model revealed that higher degree of disconnectivity was associated with higher sNfL (*t(496)* = 2.30, *β* = 0.03, *p* = 0.02), age (*t(496)* = 5.01, *β* = 0.32, *p* = 5.5 × 10^−7^) and PMS in comparison with CIS (*t(496)* = 1.97, *β* = 1.06, *p* = 0.05). The T2LV model found that higher lesions volumes were associated with increasing age (*t(496)* = 5.68, *β* = 0.22, *p* = 1.4 × 10^−8^) and timepoint (*t(496)* = 7.41, *β* = 0.02, *p* = 1.3 × 10^−13^), but not with sNfL (*t(496)* = −0.74, *β* = −0.00, *p* = 0.46). Adding centre as a random effect term in the rLMM did not affect the main results on sNfL, neither did restricting the sample to RRMS subjects only (Supplementary Table 2 and 3). We also compared model performance from the rLMM with standard linear mixed models and found converging results (Supplementary Table 4 and 5).

**Table 2.** Robust linear mixed models testing for associations of GD and T2LV with sNfL.

| *Predictors* | Global disconnectome | | | | $\beta$ | T2 lesion volume | | |
| --- | --- | --- | --- | --- | --- | --- | --- | --- |
| | $\beta$ | *CI* | *t* | *p* | | *CI* | *t* | *p* |
| (Intercept) | -1.11 | -2.16 – -0.07 | -2.09 | **0.037** | -0.66 | -1.31 – -0.01 | -1.98 | **0.047** |
| sNfL | 0.03 | 0.00 – 0.05 | 2.30 | **0.022** | -0.00 | -0.02 – 0.01 | -0.74 | 0.461 |
| Timepoint | -0.00 | -0.01 – 0.01 | -0.79 | 0.432 | 0.02 | 0.01 – 0.02 | 7.41 | **$1.3 \times 10^{-13}$** |
| Age | 0.32 | 0.19 – 0.44 | 5.01 | **$5.5 \times 10^{-7}$** | **0.22** | 0.15 – 0.30 | 5.68 | **$1.4 \times 10^{-8}$** |
| Sex [Female] | 0.11 | -0.16 – 0.37 | 0.78 | 0.434 | 0.09 | -0.08 – 0.25 | 1.01 | 0.312 |
| Diagnosis [PMS] | 1.06 | 0.01 – 2.11 | 1.97 | **0.049** | 0.38 | -0.27 – 1.04 | 1.15 | 0.249 |
| Diagnosis [RRMS] | 1.01 | -0.04 – 2.06 | 1.89 | 0.059 | 0.40 | -0.25 – 1.05 | 1.20 | 0.229 |
| Treatment [Effective] | 0.01 | -0.02 – 0.04 | 0.85 | 0.394 | -0.01 | -0.03 – 0.01 | -1.31 | 0.192 |
| Treatment [Highly-effective] | 0.01 | -0.02 – 0.04 | 0.86 | 0.389 | 0.00 | -0.01 – 0.02 | 0.46 | 0.644 |
| sNfL * Timepoint | -0.01 | -0.03 – 0.00 | -1.65 | 0.098 | 0.01 | -0.00 – 0.01 | 1.87 | 0.061 |

### Higher probability of disconnectome in cerebellum and brainstem with increased sNfL

Figure 2 shows the results from voxel-wise analysis testing for associations between disconnectome and sNfL. Briefly, the analysis revealed positive associations that were widely distributed primarily in the cerebellum and brainstem. Permutation-based corrections and TFCE revealed significant effects in the right cerebellar white matter.

**Figure 2.**
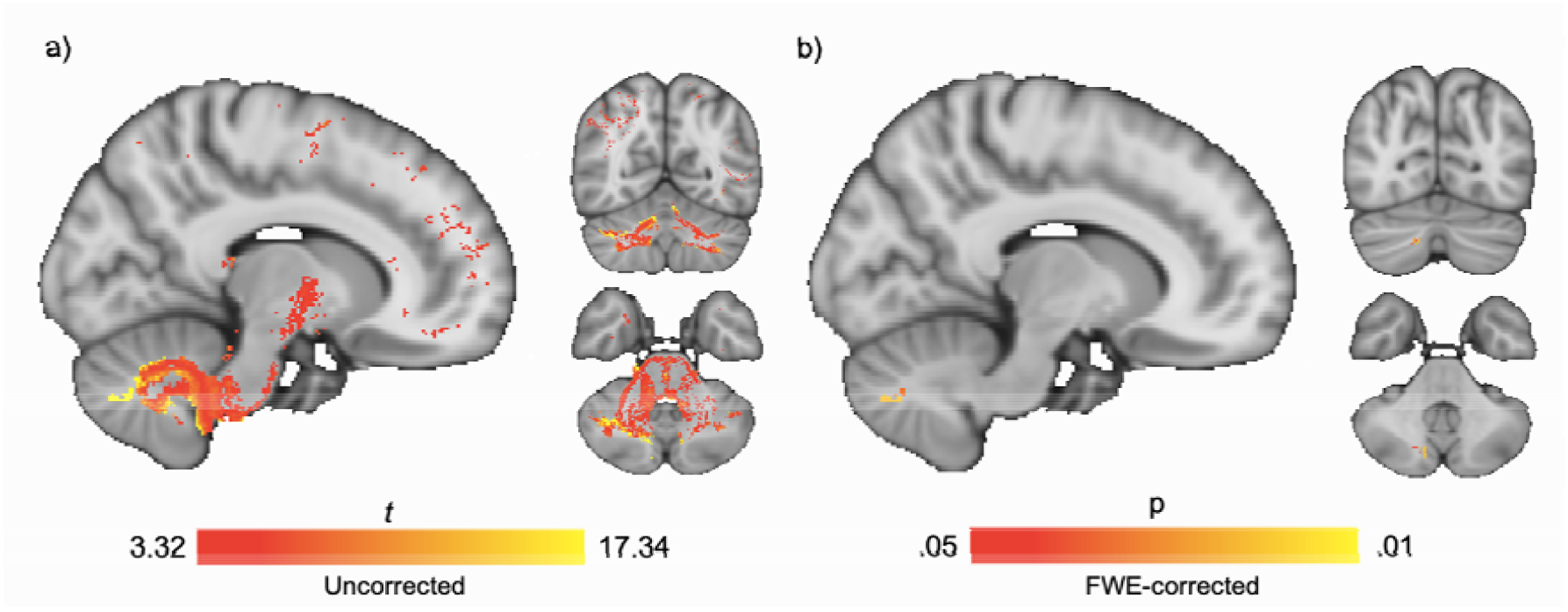
Voxel-wise analysis of disconnectome maps with increased sNfL. Regional effects of sNfL on brain disconnectome was found in cerebellum using FSL randomised with TFCE. Age, sex, diagnosis and treatment were included in the models. **(A)** Uncorrected t-stat map with threshold at *t* = 3.32, corresponding to *p* = 0.001 (uncorrected) revealed distributed associations between sNfL and disconnectome in cerebellar and brain stem regions. **(B)** FWE-corrected t-stat map with threshold at *p* = 0.05 showing a significant disconnectome cluster in cerebellum associated with increased sNfL, *t (292)* = 17.2.

## Discussion

Brain MRI is an integrated part of current multiple sclerosis clinical guidelines; it is pivotal for diagnostic and prognostic purposes, for assessing relapses and evaluating the effects of DMTs. Our understanding of the pathophysiology in multiple sclerosis is increasing, yet the clinico-radiological paradox remains unresolved.^7, 44^ Despite recent advances, MRI markers only show modest associations with clinical measures of disease progress and disability.^45^ Previous research has shown that MRI features probing physical and microstructural properties of the brain may provide sensitive markers of demyelination and axonal damage in multiple sclerosis.^44, 46^ Especially in advanced medical imaging, high-resolution diffusion MRI has been promising to evaluate brain disconnections resulting from white matter lesions ^44^. Extensive research efforts in the past two decades have supported that high central (CSF) and peripheral (plasma/serum) NfL levels reflect axonal swelling, transection and death, and may therefore represent a neuron-specific biomarker for disease processes and progression of multiple sclerosis.^21, 47^

Incorporating complex microstructural brain connectome information based on 7T diffusion imaging tractography data from healthy controls, the current disconnectome approach, utilizing T2-lesion masks to create individual brain disconnectivity maps, has provided new insights into different brain disorders,^8, 9, 34, 48^ yet the neuropathological correlates have not been investigated. Her we provide a link between imaging-based and biologically-assessed axonal damage. Our findings demonstrate that in-vivo evaluation of network-level perturbations beyond conventional T2-lesions is sensitive to neuropathological processes in multiple sclerosis, which are not necessarily identified by means of clinical neuroradiological evaluations. Expanding the initial global associations using rLMMs and voxel-wise cross-sectional analyses, revealed that the associations between the probability of disconnectivity and sNfL were primarily distributed in cerebellar and brainstem regions. While further investigations and replications are required, these findings indicate that lesions in and around white matter fibers projecting into the midbrain and cerebellum are associated with higher sNfL levels and more axonal damage. One possible explanation to our findings could be that increasing disease duration might induce more extensive damage in the cerebellar and brainstem regions, leading to an increase in NfL secretion. Supporting a potential role of cerebellum in multiple sclerosis pathophysiology and symptoms, a recent study reported that ataxia was associated with both atrophy and decreased functional connectivity in cerebellum.^49^ Furthermore, lower number of Purkinje cells has been found in demyelinated lesions in cerebellum from multiple sclerosis patients compared with healthy subjects,^50^ which could support the observed links between sNfL levels and brain disconnectivity.

The current connectivity-based approach allowed us to expand the distribution of anomalies beyond the visible T2-hyperintense brain lesions in multiple sclerosis patients, which offered a sensitive measure of neuropathology and axonal damage. Tractography and tractometry-based approaches are rare in multiple sclerosis due to technical challenges in harmonizing data from different MRI scanners and in the post-processing pipeline.^44, 51, 52^ By using the BCBtoolkit, which adapts high-resolution normative 7T diffusion data from healthy controls, we avoid complex post-processing imaging methods to incorporate brain lesions into diffusion tensor imaging (DTI) measurements. Further, since the disconnectomes are defined using tractograms from a normative and independent training set, the only required inputs are accurately defined lesion masks from each patient. This enables integration of studies employing various clinical scanning protocols and enabling large-scale, collaborative studies with the inclusion of many patient groups from which advanced MRI may not be available.

While demonstrating an association between the extent of brain disconnectivity and a neuropathological marker of axonal damage, our results have to be interpreted in the context of their limitations. The patients were enrolled from different clinical centres in a real-world setting, thus any information regarding the use of DMT are due to clinical decisions outside the scope of this paper. The mean sNfL level across all patients was relatively low, and the skewed distribution with few cases with high sNfL levels might have affected the results. Future studies may be able to test for associations among patient groups covering a larger span of disease burden and axonal damage. Further, non-random attrition e.g., due to disease progression could have resulted in a more stable longitudinal multiple sclerosis cohort. As for the normative training set, several built-in limitations and misconceptions can be introduced at all stages of the underlying tractography process.^53^ Further methodological developments and longitudinal studies are warranted to investigate the clinical and cognitive relevance and predictive value of brain disconnectome mapping in multiple sclerosis. New imaging methods incorporating the different characteristics of lesions and diffuse pathology in the normal appearing white matter could also potentially benefit this approach by more sensitive lesion characteristics.^54^

In conclusion, by providing evidence for an association between imaging-based brain disconnectome mapping and a peripheral biomarker reflecting axonal damage in patients with multiple sclerosis, these findings establish a neuropathological correlate of brain white matter affection, extending beyond conventional lesion-based characteristics. While these results support the clinical relevance of advanced network-based imaging approaches in multiple sclerosis, further studies investigating the functional and clinical sensitivity are warranted.

## Acknowledgements

We thank Ingvild Sørum Leikfoss, Ingrid Mo and Fernanda Kropf for their help with laboratory work at Oslo University Hospital, MRI technicians Susan Pikol and Cynthia Kraut for their lesion segmentations and Priscilla Bäcker-Koduah for her laboratory work at Charité-Universitätsmedizin Berlin, and all the participating multiple sclerosis patients and healthy individuals.

## Funding

This work was supported by the European Commission (ERACOSYSMED ERA-Net program, Sys4MS project, id:43), the European Union’s Horizon 2020 Research and Innovation program (ERC StG, Grant # 802998), Instituto de Salud Carlos III, Spain (AC1500052), the Italian Ministry of Health (WFR-PER-2013-02361136), the German Ministry of Science (Deutsches Teilprojekt B “Förderkennzeichen: 031L0083B), the Norwegian Research Council (257955) and Biogen Norway. Kaj Blennow is supported by the Swedish Research Council (#2017-00915), and the Swedish state under the agreement between the Swedish government and the County Councils, the ALF-agreement (#ALFGBG-715986). Michel Thiebaut de Schotten has received funding from the European Research Council (ERC) under the European Union’s Horizon 2020 research and innovation programme (grant agreement no. 818521). Henrik Zetterberg is a Wallenberg Scholar supported by grants from the Swedish Research Council (#2018-02532), the European Research Council (#681712), Swedish State Support for Clinical Research (#ALFGBG-720931), the Alzheimer Drug Discovery Foundation (ADDF), USA (#201809-2016862), the AD Strategic Fund and the Alzheimer’s Association (#ADSF-21-831376-C, #ADSF-21-831381-C and #ADSF-21-831377-C), the Olav Thon Foundation, the Erling-Persson Family Foundation, Stiftelsen för Gamla Tjänarinnor, Hjärnfonden, Sweden (#FO2019-0228), the European Union’s Horizon 2020 research and innovation programme under the Marie Skłodowska-Curie grant agreement No 860197 (MIRIADE), and the UK Dementia Research Institute at UCL.

## Competing interests

Henning H. Rise reports no disclosures.

Synne Brune has received honoraria for lecturing from Biogen and Novartis. Claudia Chien has received honoraria for lecturing from Bayer and research grants from Novartis.

Tone Berge has received unrestricted research grants from Biogen and Sanofi-Genzyme. Steffan Daniel Bos reports no disclosures.

Magi Andorrà is currently an employee of Roche, all the work in this paper is based in his previous work at IDIBAPS. He holds stock from Bionure Farma SL, Attune Neurosciences Inc and Goodgut SL.

Irene Pulido-Valdeolivas I has received travel reimbursement from Roche Spain, Novartis and Genzyme-Sanofi, and she is founder and holds stock in Aura Robotics SL. She is employee at UCB Pharma since July 2020.

Mona Beyer has received honoraria for lecturing from Novartis and Biogen Idec and served on the advisory board for Biogen.

Piotr Sowa has received honoraria for lecturing and travel support from Merck.

Michael Scheel has received funding unrelated to this work from German Research Foundation, Federal Ministry of Education and Research and Federal Ministry for Economic Affairs and Energy. He is holding patents for 3D printing of computed tomography models and is shareholder of PhantomX GmbH.

Alexander Brandt is cofounder and shareholder of Nocturne GmbH and Motognosis GmbH. He is named as inventor on several patent applications and patents describing multiple sclerosis serum biomarkers, motion analysis and retinal image analysis.

Susanna Asseyer received a conference grant from Celgene and honoraria for lecturing from Alexion, Bayer, and Roche.

Kaj Blennow has served as a consultant, at advisory boards, or at data monitoring committees for Abcam, Axon, Biogen, JOMDD/Shimadzu. Julius Clinical, Lilly, MagQu, Novartis, Roche Diagnostics, and Siemens Healthineers, and is a co-founder of Brain Biomarker Solutions in Gothenburg AB (BBS), which is a part of the GU Ventures Incubator Program.

Mads L. Pedersen reports no disclosures.

Henrik Zetterberg has served at scientific advisory boards for Eisai, Denali, Roche Diagnostics, Wave, Samumed, Siemens Healthineers, Pinteon Therapeutics, Nervgen, AZTherapies and CogRx, has given lectures in symposia sponsored by Cellectricon, Fujirebio, Alzecure and Biogen, and is a co-founder of Brain Biomarker Solutions in Gothenburg AB (BBS), which is a part of the GU Ventures Incubator Program.

Michel Thiebaut de Schotten reports no disclosures.

Marie Cellerino reports no disclosures.

Antonio Uccelli has received personal compensation from Novartis, Biogen, Merck, Roche and Sanofi Genzyme for public speaking and advisory boards. AU received funding for research by Fondazione Italiana Sclerosi Multipla, the Italian Ministry of Health and Banco San Paolo.

Friedemann Paul received honoraria and research support from Alexion, Bayer, Biogen, Chugai, MerckSerono, Novartis, Genyzme, MedImmune, Shire, Teva, and serves on scientific advisory boards for Alexion, MedImmune and Novartis. He has received funding from Deutsche Forschungsgemeinschaft (DFG Exc 257), Bundesministerium für Bildung und Forschung (Competence Network Multiple Sclerosis), Guthy Jackson Charitable Foundation, EU Framework Program 7, National Multiple Sclerosis Society of the USA.

Pablo Villoslada received consultancy fees and hold stocks from Accure Therapeutics SL, Spiral Therapeutics Inc, Clight Inc, Neuroprex Inc, QMenta Inc and Attune Neurosciences Inc.

Hanne F. Harbo has received travel support, honoraria for advice or lecturing from Biogen Idec, Sanofi-Genzyme, Merck, Novartis, Roche, and Teva and an unrestricted research grant from Novartis.

Lars T. Westlye reports no disclosures.

Einar Høgestøl received honoraria for lecturing and advisory board activity from Biogen, Merck and Sanofi-Genzyme and unrestricted research grant from Merck.

## Supplementary material

Supplementary material is available in a separate PDF file.

## Appendix 1

List of all authors and their individual contribution to this manuscript.

| Name | Location | Role | Contribution |
| --- | --- | --- | --- |
| H. H. Rise, MSc | Division of Mental Health and Addiction, Oslo | First Author | Conception and design of the work; data analysis and interpretation; drafting the article; critical revision of the article; final approval of the version to be published |
| S. Brune, MD | University of Oslo | Author | Conception of the work; data collection; data interpretation; drafting the article; critical revision of the article; final approval of the version to be published |
| Claudia Chien, PhD | Charité-Universitätsmedizin Berlin | Author | Data collection; data analysis (MRI pre-processing) and interpretation; drafting the article; critical revision of the article; final approval of the version to be published |
| T. Berge, PhD | University of Oslo | Author | Conception of the work; data collection; data interpretation; drafting the article; critical revision of the article; final approval of the version to be published |
| S.D. Bos, PhD | University of Oslo | Author | Data collection; data interpretation; critical revision of the article; final approval of the version to be published |
| Magi Andorra, PhD | IDIBAPS | Author | Data collection; critical revision of the article, final approval of the version to be published |
| Irene Pulido<br>Valdeolivas, MD | IDIBAPS | Author | Data collection; critical revision of the article, final approval of the version to be published |
| M. K. Beyer, MD, PhD | Oslo University Hospital | Author | Data collection; critical revision of the article, final approval of the version to be published |
| P. Sowa, MD, PhD | Oslo University Hospital | Author | Data collection; critical revision of the article, final approval of the version to be published |
| Michael Scheel, MD | Charité<br>Universitaetsmedizin<br>Berlin | Author | Data collection; Data analysis (MRI); critical revision of the article, final approval of the version to be published |
| Alexander Brandt, MD, PhD | Charité<br>Universitaetsmedizin<br>Berlin | Author | Data collection; Data analysis (MRI); critical revision of the article, final approval of the version to be published |
| Susanna Asseyer, MD | Charité<br>Universitaetsmedizin<br>Berlin | Author | Data collection; critical revision of the article, final approval of the version to be published |
| K. Blennow, PhD | University of Gothenburg | Author | Data collection (laboratory analyses), critical revision of the article, final approval of the version to be published |
| Mads L. Pedersen, PhD | University of Oslo | Author | Data analysis (linear mixed models) and interpretation; critical revision of the article; final approval of the version to be published |
| Henrik Zetterberg,<br>PhD | University of Gothenburg | Author | Data collection (laboratory analyses), critical revision of the article, final approval of the version to be published |
| Michel Thiebaut de<br>Schotten, PhD | CNRS | Author | Data analysis (disconnectome 7T), Revision of the article & final approval of the version to be published |
| Marie Cellerino,<br>PhD | University of Genoa | Author | Data collection; Revision of the article & final approval of the version to be published |
| A. Uccelli, MD, PhD | University of Genoa | Author | Design and conceptualized study; contributed to the design of the study and revised the manuscript for intellectual content |
| F. Paul, MD, PhD | Charité<br>Universitätsmedizin<br>Berlin | Author | Design and conceptualized study; contributed to the design of the study and revised the manuscript for intellectual content |
| Pablo Villoslada,<br>MD, PhD | IDIBAPS | Author | Design and conceptualized study; contributed to the design of the study and revised the manuscript for intellectual content |
| H. F. Harbo, MD,<br>PhD | University of Oslo and<br>Oslo University Hospital | Author | Design and conceptualized study; contributed to the design of the study and revised the manuscript for intellectual content |
| L. T. Westlye, PhD | University of Oslo and<br>Oslo University Hospital | Author | Design and conceptualized study; interpreted the data; drafting the article; revised the manuscript for intellectual content |
| E. A. Høgestøl, MD,<br>PhD | Oslo University Hospital | Last author<br>(Corresponding author) | Design and conceptualized study; data collection; data interpretation; drafting the article; interpreted the data; revised the manuscript for intellectual content |

## Abbreviations

3D BRAVO: 3D sagittal brain volume
CAT12: Computational Anatomy Toolbox for SPM12
CIS: clinically isolated syndrome
DMT: disease modifying treatments
DTI: diffusion tensor imaging
EDSS: expanded disability status scale
FLAIR: fluid-attenuated inversion recovery
FSPGR: fast-spoiled gradient-echo
GA: glatiramer acetate
GD: global disconnectivity
GLM: general linear model
HC: healthy controls
ID: identifier
IFN: interferon
IQR: Interquartile range
MNI: Montreal Neurological Institute
MPRAGE: magnetization prepared rapid gradient echo
MRI: magnetic resonance imaging, alphabetical order
NfL: neurofilament light chain
PBVC: percent brain tissue volume change
PMS: progressive multiple sclerosis
RIS: radiologically isolated syndrome
rLMM: robust linear mixed-effects models
RRMS: remitting-remitting multiple sclerosis
SIMOA: single molecule array
T: Tesla
T2LV: T2 lesion volume
TE: echo time
TFCE: threshold-free cluster enhancement
TFE: turbo field echo
TR: repetition time

## References

1. Dobson R, Giovannoni G. Multiple sclerosis - a review. Eur J Neurol. Jan 2019;26(1):27–40. doi:10.1111/ene.13819

2. Thompson AJ, Baranzini SE, Geurts J, Hemmer B, Ciccarelli O. Multiple sclerosis. Lancet. Apr 21 2018;391(10130):1622–1636. doi:10.1016/S0140-6736(18)30481-1

3. Geraldes R, Ciccarelli O, Barkhof F, et al. The current role of MRI in differentiating multiple sclerosis from its imaging mimics. Nature reviews Neurology. Mar 9 2018;doi:10.1038/nrneurol.2018.14

4. Filippi M, Rocca MA, Ciccarelli O, et al. MRI criteria for the diagnosis of multiple sclerosis: MAGNIMS consensus guidelines. Lancet Neurol. Mar 2016;15(3):292–303. doi:10.1016/s1474-4422(15)00393-2

5. Filippi M, Preziosa P, Banwell BL, et al. Assessment of lesions on magnetic resonance imaging in multiple sclerosis: practical guidelines. Brain. Jun 17 2019;doi:10.1093/brain/awz144

6. Barkhof F. The clinico-radiological paradox in multiple sclerosis revisited. Curr Opin Neurol. Jun 2002;15(3):239–45. doi:10.1097/00019052-200206000-00003

7. Mollison D, Sellar R, Bastin M, et al. The clinico-radiological paradox of cognitive function and MRI burden of white matter lesions in people with multiple sclerosis: A systematic review and meta-analysis. PLoS One. 2017;12(5):e0177727. doi:10.1371/journal.pone.0177727

8. Thiebaut de Schotten M, Foulon C, Nachev P. Brain disconnections link structural connectivity with function and behaviour. Nat Commun. Oct 9 2020;11(1):5094. doi:10.1038/s41467-020-18920-9

9. Thiebaut de Schotten M, Dell’Acqua F, Ratiu P, et al. From Phineas Gage and Monsieur Leborgne to H.M.: Revisiting Disconnection Syndromes. Cereb Cortex. Dec 2015;25(12):4812–27. doi:10.1093/cercor/bhv173

10. Gaetani L, Blennow K, Calabresi P, Di Filippo M, Parnetti L, Zetterberg H. Neurofilament light chain as a biomarker in neurological disorders. J Neurol Neurosurg Psychiatry. 2019;90(8):870–881. doi:10.1136/jnnp-2018-320106

11. Kuhle J, Barro C, Andreasson U, et al. Comparison of three analytical platforms for quantification of the neurofilament light chain in blood samples: ELISA, electrochemiluminescence immunoassay and Simoa. Clin Chem Lab Med. Oct 1 2016;54(10):1655–61. doi:10.1515/cclm-2015-1195

12. Disanto G, Barro C, Benkert P, et al. Serum Neurofilament light: A biomarker of neuronal damage in multiple sclerosis. Annals of neurology. Jun 2017;81(6):857–870. doi:10.1002/ana.24954

13. Lycke JN, Karlsson JE, Andersen O, Rosengren LE. Neurofilament protein in cerebrospinal fluid: a potential marker of activity in multiple sclerosis. J Neurol Neurosurg Psychiatry. Mar 1998;64(3):402–4. doi:10.1136/jnnp.64.3.402

14. Tavazzi E, Jakimovski D, Kuhle J, et al. Serum neurofilament light chain and optical coherence tomography measures in MS: A longitudinal study. Neurol Neuroimmunol Neuroinflamm. Jul 2020;7(4)doi:10.1212/NXI.0000000000000737

15. Malmeström C, Haghighi S, Rosengren L, Andersen O, Lycke J. Neurofilament light protein and glial fibrillary acidic protein as biological markers in MS. Neurology. Dec 23 2003;61(12):1720–5. doi:10.1212/01.wnl.0000098880.19793.b6

16. Gunnarsson M, Malmeström C, Axelsson M, et al. Axonal damage in relapsing multiple sclerosis is markedly reduced by natalizumab. Annals of neurology. Jan 2011;69(1):83–9. doi:10.1002/ana.22247

17. Piehl F, Kockum I, Khademi M, et al. Plasma neurofilament light chain levels in patients with MS switching from injectable therapies to fingolimod. Multiple sclerosis (Houndmills, Basingstoke, England). Jul 2018;24(8):1046–1054. doi:10.1177/1352458517715132

18. Hakansson I, Tisell A, Cassel P, et al. Neurofilament light chain in cerebrospinal fluid and prediction of disease activity in clinically isolated syndrome and relapsing-remitting multiple sclerosis. Eur J Neurol. May 2017;24(5):703–712. doi:10.1111/ene.13274

19. Barro C, Benkert P, Disanto G, et al. Serum neurofilament as a predictor of disease worsening and brain and spinal cord atrophy in multiple sclerosis. Brain. Aug 1 2018;141(8):2382–2391. doi:10.1093/brain/awy154

20. Varhaug KN, Barro C, Bjornevik K, et al. Neurofilament light chain predicts disease activity in relapsing-remitting MS. Neurol Neuroimmunol Neuroinflamm. Jan 2018;5(1):e422. doi:10.1212/NXI.0000000000000422

21. Preziosa P, Rocca MA, Filippi M. Current state-of-art of the application of serum neurofilaments in multiple sclerosis diagnosis and monitoring. Expert Rev Neurother. Apr 23 2020;doi:10.1080/14737175.2020.1760846

22. Ineichen BV, Moridi T, Ewing E, et al. Neurofilament light chain as a marker for cortical atrophy in multiple sclerosis without radiological signs of disease activity. J Intern Med. n/a(n/a)doi:https://doi.org/10.1111/joim.13286

23. Bjornevik K, Munger KL, Cortese M, et al. Serum Neurofilament Light Chain Levels in Patients With Presymptomatic Multiple Sclerosis. JAMA neurology. Jan 1 2020;77(1):58–64. doi:10.1001/jamaneurol.2019.3238

24. Joisten N, Rademacher A, Warnke C, et al. Exercise Diminishes Plasma Neurofilament Light Chain and Reroutes the Kynurenine Pathway in Multiple Sclerosis. Neurol Neuroimmunol Neuroinflamm. May 2021;8(3)doi:10.1212/NXI.0000000000000982

25. Saraste M, Bezukladova S, Matilainen M, et al. High serum neurofilament associates with diffuse white matter damage in MS. Neurol Neuroimmunol Neuroinflamm. Jan 2021;8(1)doi:10.1212/NXI.0000000000000926

26. Bridel C, Verberk IMW, Heijst JJA, Killestein J, Teunissen CE. Variations in consecutive serum neurofilament light levels in healthy controls and multiple sclerosis patients. Multiple sclerosis and related disorders. Jan 2021;47:102666. doi:10.1016/j.msard.2020.102666

27. Polman CH, Reingold SC, Banwell B, et al. Diagnostic criteria for multiple sclerosis: 2010 revisions to the McDonald criteria. Annals of neurology. Feb 2011;69(2):292–302. doi:10.1002/ana.22366

28. Ashburner J, Friston KJ. Unified segmentation. Neuroimage. Jul 1 2005;26(3):839–51. doi:10.1016/j.neuroimage.2005.02.018

29. Jenkinson M, Smith S. A global optimisation method for robust affine registration of brain images. Med Image Anal. Jun 2001;5(2):143–56. doi:10.1016/s1361-8415(01)00036-6

30. Jenkinson M, Bannister P, Brady M, Smith S. Improved optimization for the robust and accurate linear registration and motion correction of brain images. Neuroimage. Oct 2002;17(2):825–41.

31. Yushkevich PA, Piven J, Hazlett HC, et al. User-guided 3D active contour segmentation of anatomical structures: significantly improved efficiency and reliability. Neuroimage. Jul 1 2006;31(3):1116–28. doi:10.1016/j.neuroimage.2006.01.015

32. Smith SM, Zhang Y, Jenkinson M, et al. Accurate, robust, and automated longitudinal and cross-sectional brain change analysis. Neuroimage. Sep 2002;17(1):479–89.

33. Smith SM, Jenkinson M, Woolrich MW, et al. Advances in functional and structural MR image analysis and implementation as FSL. Neuroimage. 2004;23 Suppl 1:S208–19. doi:10.1016/j.neuroimage.2004.07.051

34. Foulon C, Cerliani L, Kinkingnehun S, et al. Advanced lesion symptom mapping analyses and implementation as BCBtoolkit. Gigascience. Mar 1 2018;7(3):1–17. doi:10.1093/gigascience/giy004

35. Rojkova K, Volle E, Urbanski M, Humbert F, Dell’Acqua F, Thiebaut de Schotten M. Atlasing the frontal lobe connections and their variability due to age and education: a spherical deconvolution tractography study. Brain Struct Funct. Apr 2016;221(3):1751–66. doi:10.1007/s00429-015-1001-3

36. Thiebaut de Schotten M, Dell’Acqua F, Forkel SJ, et al. A lateralized brain network for visuospatial attention. Nat Neurosci. Sep 18 2011;14(10):1245–6. doi:10.1038/nn.2905

37. Klein A, Andersson J, Ardekani BA, et al. Evaluation of 14 nonlinear deformation algorithms applied to human brain MRI registration. Neuroimage. Jul 1 2009;46(3):786–802. doi:10.1016/j.neuroimage.2008.12.037

38. Avants BB, Tustison NJ, Song G, Cook PA, Klein A, Gee JC. A reproducible evaluation of ANTs similarity metric performance in brain image registration. Neuroimage. Feb 1 2011;54(3):2033–44. doi:10.1016/j.neuroimage.2010.09.025

39. R. Wang TB, A. G. Sorensen, and V. J. Wedeen. Diffusion Toolkit: A Software Package for Diffusion Imaging Data Processing and Tractography. https://cds.ismrm.org/ismrm-2007/files/03720.pdf: Martinos Center for Biomedical Imaging, MGH, Charlestown, MA, United States; 2007.

40. Winkler AM, Ridgway GR, Webster MA, Smith SM, Nichols TE. Permutation inference for the general linear model. Neuroimage. May 15 2014;92:381–97. doi:10.1016/j.neuroimage.2014.01.060

41. Smith SM, Nichols TE. Threshold-free cluster enhancement: addressing problems of smoothing, threshold dependence and localisation in cluster inference. Neuroimage. Jan 1 2009;44(1):83–98. doi:10.1016/j.neuroimage.2008.03.061

42. R: A Language and Environment for Statistical Computing. R Foundation for Statistical Computing; 2017. https://www.R-project.org/

43. Koller M. robustlmm: An R Package for Robust Estimation of Linear Mixed-Effects Models. robust statistics; mixed-effects model; hierarchical model; ANOVA; R; crossed; random effect. 2016. 2016-12-06 2016;75(6):24. doi:10.18637/jss.v075.i06

44. Chamberland M, Winter M, Brice T, Jones D, Tallantyre E. Beyond lesion-load: Tractometry-based metrics for characterizing white matter lesions within fibre pathways. 2020.

45. Filippi M, Preziosa P, Langdon D, et al. Identifying Progression In Multiple Sclerosis: New Perspectives. Annals of neurology. Jun 7 2020;doi:10.1002/ana.25808

46. Rocca MA, Amato MP, De Stefano N, et al. Clinical and imaging assessment of cognitive dysfunction in multiple sclerosis. Lancet Neurol. Mar 2015;14(3):302–17. doi:10.1016/s1474-4422(14)70250-9

47. Khalil M, Pirpamer L, Hofer E, et al. Serum neurofilament light levels in normal aging and their association with morphologic brain changes. Nat Commun. Feb 10 2020;11(1):812. doi:10.1038/s41467-020-14612-6

48. Ulrichsen KM, Kolskår KK, Richard G, et al. Structural brain disconnectivity mapping of post-stroke fatigue. NeuroImage: Clinical. 2021/03/22/ 2021:102635. doi:https://doi.org/10.1016/j.nicl.2021.102635

49. Cordani C, Meani A, Esposito F, et al. Imaging correlates of hand motor performance in multiple sclerosis: A multiparametric structural and functional MRI study. Multiple sclerosis (Houndmills, Basingstoke, England). Feb 2020;26(2):233–244. doi:10.1177/1352458518822145

50. Redondo J, Kemp K, Hares K, Rice C, Scolding N, Wilkins A. Purkinje Cell Pathology and Loss in Multiple Sclerosis Cerebellum. Brain Pathol. Nov 2015;25(6):692–700. doi:10.1111/bpa.12230

51. Ciccarelli O, Catani M, Johansen-Berg H, Clark C, Thompson A. Diffusion-based tractography in neurological disorders: concepts, applications, and future developments. Lancet Neurol. Aug 2008;7(8):715–27. doi:10.1016/S1474-4422(08)70163-7

52. Lipp I, Parker GD, Tallantyre EC, et al. Tractography in the presence of multiple sclerosis lesions. Neuroimage. Apr 1 2020;209:116471. doi:10.1016/j.neuroimage.2019.116471

53. Schilling KG, Nath V, Hansen C, et al. Limits to anatomical accuracy of diffusion tractography using modern approaches. Neuroimage. Jan 15 2019;185:1–11. doi:10.1016/j.neuroimage.2018.10.029

54. Chard DT, Alahmadi AAS, Audoin B, et al. Mind the gap: from neurons to networks to outcomes in multiple sclerosis. Nature reviews Neurology. Jan 12 2021;doi:10.1038/s41582-020-00439-8

