## Supplementary for "Brain disconnectome mapping and serum neurofilament light levels in multiple sclerosis"

### Supplementary material

**Supplementary Table 1 Demographic characteristics and sNfL in the healthy control group**

| | Baseline | Follow-up |
|---|---|---|
| | n = 59 | n = 30 |
| Female % (n) | 78 (47) | 83 (25) |
| Age, mean years (SD, range) | 39.9 (11.8, 22-71) | 41.2 (12.5, 26-73) |
| Serum Neurofilament Light levels, mean pgmL (SD, range) | 7.0 (3.8, 2.2-21.6) | 7.8 (3.7, 3.0-16.2) |

*SD, standard deviation*

#### Sensitivity analysis

To explore the effects of centre in our rLMM models we added center as a random effect term (Supplementary Table 2). In addition, we also used the same set-up with centre while restricting the sample to RRMS subjects only (Supplementary Table 3).

**Supplementary Table 2 Robust linear mixed models predicting sNfL with global disconnectome and lesion volume including centre as random effect term**

| | GD | | | | T2LV | | | |
|---|---|---|---|---|---|---|---|---|
| *Predictors* | *Estimates* | *CI* | *t* | *p* | *Estimates* | *CI* | *t* | *p* |
| (Intercept) | -0.79 | -1.87 – 0.29 | -1.44 | 0.151 | -0.51 | -1.19 – 0.16 | -1.48 | 0.138 |
| sNfL | 0.03 | 0.01 – 0.05 | 2.94 | **0.003** | -0.00 | -0.02 – 0.01 | -0.76 | 0.450 |
| Timepoint | -0.00 | -0.01 – 0.00 | -0.88 | 0.377 | 0.02 | 0.01 – 0.02 | 7.45 | **<0.001** |
| Age | 0.26 | 0.14 – 0.37 | 4.36 | **<0.001** | 0.20 | 0.13 – 0.27 | 5.43 | **<0.001** |
| Sex [Female] | 0.07 | -0.18 – 0.32 | 0.58 | 0.560 | 0.08 | -0.08 – 0.23 | 0.96 | 0.339 |
| diagnosis [PMS] | 0.72 | -0.27 – 1.70 | 1.42 | 0.155 | 0.22 | -0.40 – 0.84 | 0.69 | 0.492 |
| diagnosis [RRMS] | 0.67 | -0.31 – 1.66 | 1.34 | 0.180 | 0.24 | -0.38 – 0.86 | 0.75 | 0.453 |
| treatment [Effective] | 0.01 | -0.02 – 0.04 | 0.83 | 0.406 | -0.01 | -0.03 – 0.00 | -1.35 | 0.178 |
| treatment [Highly-effective] | 0.01 | -0.02 – 0.04 | 0.81 | 0.417 | 0.00 | -0.01 – 0.02 | 0.36 | 0.720 |

| | | | | | | | | |
|---|---|---|---|---|---|---|---|---|
| sNfL * Timepoint | -0.01 | -0.03 – -0.00 | -2.31 | **0.021** | 0.01 | -0.00 – 0.01 | 1.92 | 0.055 |
| **Random Effects** | | | | | | | | |
| $\sigma^2$ | 0.00 | | | | 0.00 | | | |
| $\tau_{00}$ | 0.89 ID | | | | 0.35 ID | | | |
| | 0.19 center | | | | 0.07 center | | | |
| ICC | 1.00 | | | | 1.00 | | | |
| N | 296 ID | | | | 296 ID | | | |
| | 4 center | | | | 4 center | | | |
| Observations | 507 | | | | 506 | | | |
| Marginal $R^2$ / Conditional $R^2$ | 0.070 / 0.998 | | | | 0.090 / 0.999 | | | |

**Supplementary Table 3 Robust linear mixed models predicting sNfL with global disconnectome and lesion volume including centre as random effect term for RRMS subjects only**

| | GD | | | | T2LV | | | |
|---|---|---|---|---|---|---|---|---|
| *Predictors* | *Estimates* | *CI* | *t* | *p* | *Estimates* | *CI* | *t* | *p* |
| (Intercept) | -0.14 | -0.64 – 0.37 | -0.53 | 0.595 | -0.30 | -0.59 – -0.00 | -1.98 | **0.048** |
| sNfL | 0.03 | 0.01 – 0.06 | 2.66 | **0.008** | 0.00 | -0.02 – 0.02 | 0.03 | 0.976 |
| Timepoint | -0.00 | -0.01 – 0.01 | -0.40 | 0.692 | 0.02 | 0.01 – 0.03 | 6.09 | **<0.001** |
| Age | 0.17 | 0.04 – 0.31 | 2.62 | **0.009** | 0.12 | 0.04 – 0.20 | 2.98 | **0.003** |
| Sex [Female] | 0.09 | -0.20 – 0.39 | 0.62 | 0.535 | 0.12 | -0.06 – 0.29 | 1.27 | 0.204 |
| treatment [Effective] | 0.03 | -0.01 – 0.06 | 1.43 | 0.152 | -0.00 | -0.02 – 0.02 | -0.10 | 0.923 |
| treatment [Highly-effective] | 0.05 | 0.01 – 0.08 | 2.32 | **0.021** | 0.03 | 0.01 – 0.05 | 2.50 | **0.012** |
| sNfL * Timepoint | -0.02 | -0.03 – -0.00 | -2.20 | **0.028** | 0.00 | -0.01 – 0.02 | 0.55 | 0.580 |
| **Random Effects** | | | | | | | | |
| $\sigma^2$ | 0.00 | | | | 0.00 | | | |
| $\tau_{00}$ | 0.99 ID | | | | 0.35 ID | | | |
| | 0.18 center | | | | 0.06 center | | | |
| ICC | 1.00 | | | | 1.00 | | | |
| N | 243 ID | | | | 243 ID | | | |
| | 4 center | | | | 4 center | | | |

| | | |
|---|---|---|
| Observations | 412 | 411 |
| Marginal $R^2$ / Conditional $R^2$ | 0.027 / 0.998 | 0.039 / 0.998 |

#### Comparing statistical output

We also performed comparing analyses comparing the output from more regular linear mixed models (LMM) with the robust linear mixed models that we performed as our main analysis. For global disconnectome (GD) see Supplementary Table 4, and for T2 lesion volume see Supplementary Table 5.

**Supplementary Table 4 Overview of the model performance of linear mixed models compared with robust linear mixed models for global disconnectome.**

| | Linear mixed models | | | | Robust linear mixed models | | | |
|---|---|---|---|---|---|---|---|---|
| | sNfL | | | | sNfL | | | |
| *Predictors* | *Estimates* | *CI* | *Statistic* | *p* | *Estimates* | *CI* | *Statistic* | *p* |
| (Intercept) | -0.27 | -1.25 – 0.72 | -0.53 | 0.597 | -0.29 | -0.72 – 0.13 | -1.34 | 0.179 |
| GD | 0.18 | 0.01 – 0.35 | 2.06 | **0.039** | 0.09 | 0.01 – 0.17 | 2.27 | **0.023** |
| GD * Timepoint | 0.01 | -0.08 – 0.10 | 0.25 | 0.801 | -0.01 | -0.06 – 0.03 | -0.61 | 0.542 |
| Timepoint | 0.00 | -0.09 – 0.09 | 0.06 | 0.950 | 0.01 | -0.04 – 0.05 | 0.27 | 0.790 |
| Age | 0.02 | -0.12 – 0.15 | 0.23 | 0.819 | 0.16 | 0.11 – 0.22 | 5.51 | **<0.001** |
| Sex [Female] | 0.14 | -0.11 – 0.39 | 1.09 | 0.276 | 0.08 | -0.03 – 0.18 | 1.38 | 0.169 |
| Diagnosis [PMS] | 0.64 | -0.39 – 1.68 | 1.22 | 0.223 | 0.37 | -0.08 – 0.81 | 1.61 | 0.107 |
| Diagnosis [RRMS] | 0.22 | -0.76 – 1.21 | 0.44 | 0.658 | 0.13 | -0.29 – 0.56 | 0.61 | 0.541 |
| Treatment [Effective] | -0.09 | -0.32 – 0.13 | -0.81 | 0.416 | -0.10 | -0.21 – -0.00 | -1.99 | **0.046** |
| Treatment [Highly-effective] | -0.25 | -0.47 – -0.03 | -2.21 | **0.027** | -0.12 | -0.23 – -0.02 | -2.34 | **0.019** |
| **Random Effects** | | | | | | | | |
| $\sigma^2$ | 0.23 | | | | 0.05 | | | |
| $\tau_{00}$ | 0.80 ID | | | | 0.13 ID | | | |
| ICC | 0.78 | | | | 0.71 | | | |
| N | 296 ID | | | | 296 ID | | | |
| Observations | 507 | | | | 507 | | | |
| Marginal $R^2$ / Conditional $R^2$ | 0.094 / 0.799 | | | | 0.295 / 0.799 | | | |

**Supplementary Table 5 Overview of the model performance of linear mixed models compared with robust linear mixed models for T2 lesion volume.**

| | Linear mixed models | | | | Robust linear mixed models | | | |
|---|---|---|---|---|---|---|---|---|
| | sNfL | | | | sNfL | | | |
| *Predictors* | *Estimates* | *CI* | *Statistic* | *p* | *Estimates* | *CI* | *Statistic* | *p* |
| (Intercept) | -0.43 | -1.40 – 0.55 | -0.85 | 0.393 | -0.35 | -0.77 – 0.07 | -1.62 | 0.106 |
| T2LV | 0.08 | -0.10 – 0.25 | 0.89 | 0.374 | 0.09 | 0.01 – 0.17 | 2.11 | **0.035** |
| T2LV * Timepoint | 0.01 | -0.08 – 0.10 | 0.20 | 0.842 | -0.03 | -0.07 – 0.02 | -1.27 | 0.205 |
| Timepoint | -0.01 | -0.10 – 0.09 | -0.17 | 0.867 | 0.01 | -0.04 – 0.05 | 0.22 | 0.823 |
| Age | 0.04 | -0.09 – 0.17 | 0.58 | 0.563 | 0.17 | 0.11 – 0.23 | 5.79 | **<0.001** |
| Sex [Female] | 0.15 | -0.09 – 0.40 | 1.22 | 0.224 | 0.08 | -0.03 – 0.19 | 1.43 | 0.152 |
| Diagnosis [PMS] | 0.83 | -0.20 – 1.86 | 1.58 | 0.114 | 0.43 | -0.02 – 0.87 | 1.89 | 0.059 |
| Diagnosis [RRMS] | 0.35 | -0.63 – 1.33 | 0.70 | 0.485 | 0.17 | -0.25 – 0.59 | 0.80 | 0.421 |
| Treatment [Effective] | -0.06 | -0.29 – 0.16 | -0.56 | 0.576 | -0.09 | -0.19 – 0.01 | -1.79 | 0.074 |
| Treatment [Highly-effective] | -0.21 | -0.43 – 0.02 | -1.81 | 0.071 | -0.11 | -0.21 – -0.01 | -2.08 | **0.037** |
| **Random Effects** | | | | | | | | |
| $\sigma^2$ | 0.24 | | | | 0.05 | | | |
| $\tau_{00}$ | 0.79 $_{ID}$ | | | | 0.13 $_{ID}$ | | | |
| ICC | 0.77 | | | | 0.71 | | | |
| N | 296 $_{ID}$ | | | | 296 $_{ID}$ | | | |
| Observations | | | | 506 | | | | 506 |
| Marginal $R^2$ / Conditional $R^2$ | 0.076 / 0.786 | | | | 0.294 / 0.797 | | | |

#### Outlier analysis

Outlier detection for sNfL scores was performed following Tukey's fence method, where a score is considered an outlier if the value is either below the first quartile - 1.5 * interquartile range (IQR) or above the third quartile + 1.5* IQR. However, removal of sNfL outliers did not affect the overall results, as described in Supplementary Table 6.

**Supplementary Table 6 Robust linear mixed models predicting sNfL with global disconnectome and lesion volume after removing sNfL outliers**

| | GD | | | | T2LV | | | |
|---|---|---|---|---|---|---|---|---|
| *Predictors* | *Estimates* | *CI* | *t* | *p* | *Estimates* | *CI* | *t* | *p* |
| (Intercept) | -1.22 | -2.22 – -0.21 | -2.38 | **0.017** | -0.75 | -1.36 – -0.13 | -2.39 | **0.017** |
| sNfL | 0.02 | 0.00 – 0.04 | 2.19 | **0.028** | -0.00 | -0.01 – 0.01 | -0.29 | 0.770 |
| Timepoint | -0.00 | -0.01 – 0.01 | -0.49 | 0.622 | 0.02 | 0.01 – 0.02 | 7.03 | **$2.0x10^{-12}$** |
| Age | 0.20 | 0.06 – 0.34 | 2.76 | **0.006** | 0.13 | 0.04 – 0.22 | 2.99 | **0.003** |
| Sex [Female] | 0.14 | -0.12 – 0.40 | 1.06 | 0.290 | 0.13 | -0.03 – 0.29 | 1.58 | 0.113 |
| Diagnosis [PMS] | 1.73 | 0.67 – 2.80 | 3.19 | **0.001** | 1.17 | 0.52 – 1.82 | 3.54 | **$4.1x10^{-4}$** |
| Diagnosis [RRMS] | 0.96 | -0.05 – 1.96 | 1.87 | 0.061 | 0.35 | -0.26 – 0.97 | 1.14 | 0.256 |
| Treatment [Effective] | 0.01 | -0.02 – 0.04 | 0.50 | 0.616 | -0.01 | -0.02 – 0.01 | -0.90 | 0.369 |
| Treatment [Highly-effective] | 0.01 | -0.02 – 0.04 | 0.66 | 0.507 | -0.00 | -0.02 – 0.01 | -0.17 | 0.869 |
| sNfL * Timepoint | -0.01 | -0.02 – 0.00 | -1.19 | 0.233 | 0.00 | -0.00 – 0.01 | 1.74 | 0.082 |
| **Random Effects** | | | | | | | | |
| $\sigma^2$ | 0.00 | | | | 0.00 | | | |
| $\tau_{00}$ | 0.97 $_{ID}$ | | | | 0.36 $_{ID}$ | | | |
| ICC | 1.00 | | | | 1.00 | | | |
| N | 287 $_{ID}$ | | | | 287 $_{ID}$ | | | |
| Observations | 484 | | | | 484 | | | |
| Marginal $R^2$ | 0.169 | | | | 0.294 | | | |

Outliers identified: 23 from 512 observations across both timepoint
Proportion (%) of outliers: 4.49
Mean of the outliers: 28.74
Mean sNfL without removing outliers: 8.80
Mean sNfL after removing outliers: 7.86

### Cross-sectional analyses with multiple linear regression models

To investigate associations between sNfL and GD and T2LV at baseline, two separate multiple linear models where conducted, with GD and T2LV as dependent variables, respectively.

Supplementary Table 7 summarizes the results from linear models testing for associations between GD and T2LV with sNfL levels, different treatments and MS phenotypes at baseline. Briefly, the model revealed significant associations between sNfL and GD (t(286) = 4.62, p < .001), age (t(286) = 3.90, p < .001), and diagnosis (t(286) = 3.94, p < .001), indicating higher level of dysconnectivity with higher NfL, higher age and with PMS compared to CIS subtype. Significant effects were also evident for both DMT groups compared to no treatment, with effective treatment (t(286) = 3.29, p = .001) and highly-effective treatment (t(286) = 4.75, p < .001) being associated with higher levels of brain dysconnectivity. The T2LV models revealed a significant association with sNfL (t(286) = 2.89, p = .004). In addition, the use of any DMTs compared to no treatment was associated with larger lesions, for both effective treatment (t(286) = 2.71, p = .007), as well as highly-effective treatment (t(286) = 3.49, p = .001).

**Supplementary Table 7 Linear regression for global disconnectome and lesion volume at baseline with sNfL**

| | GD | | | | T2LV | | | |
|---|---|---|---|---|---|---|---|---|
| *Predictors* | *Estimates* | *CI* | *t* | *p* | *Estimates* | *CI* | *t* | *p* |
| (Intercept) | -1.16 | -1.77 – -0.55 | -3.70 | **$2.6x10^{-4}$** | -0.70 | -0.83 – -0.56 | -10.29 | **$1.5x10^{-21}$** |
| sNfL | 0.14 | 0.08 – 0.21 | 4.62 | **$5.7x10^{-6}$** | 0.05 | 0.02 – 0.09 | 2.89 | **0.004** |
| Age | 0.24 | 0.12 – 0.36 | 3.90 | **$1.2x10^{-4}$** | 0.08 | -0.01 – 0.17 | 1.68 | 0.094 |
| Sex [Female] | 0.13 | -0.08 – 0.34 | 1.19 | 0.234 | 0.03 | -0.08 – 0.14 | 0.54 | 0.592 |
| Diagnosis [PMS] | 1.39 | 0.70 – 2.09 | 3.94 | **$1.0x10^{-4}$** | 0.37 | -0.00 – 0.74 | 1.95 | 0.052 |
| Diagnosis [RRMS] | 0.52 | -0.11 – 1.15 | 1.63 | 0.104 | 0.11 | -0.03 – 0.26 | 1.56 | 0.121 |
| Treatment [Effective] | 0.44 | 0.18 – 0.70 | 3.29 | **0.001** | 0.17 | 0.05 – 0.29 | 2.71 | **0.007** |
| Treatment [Highly-effective] | 0.73 | 0.43 – 1.04 | 4.75 | **$3.3x10^{-6}$** | 0.35 | 0.15 – 0.55 | 3.49 | **0.001** |
| Observations | 294 | | | | 294 | | | |
| $R^2$ / $R^2$ adjusted | 0.261 / 0.243 | | | | 0.160 / 0.139 | | | |

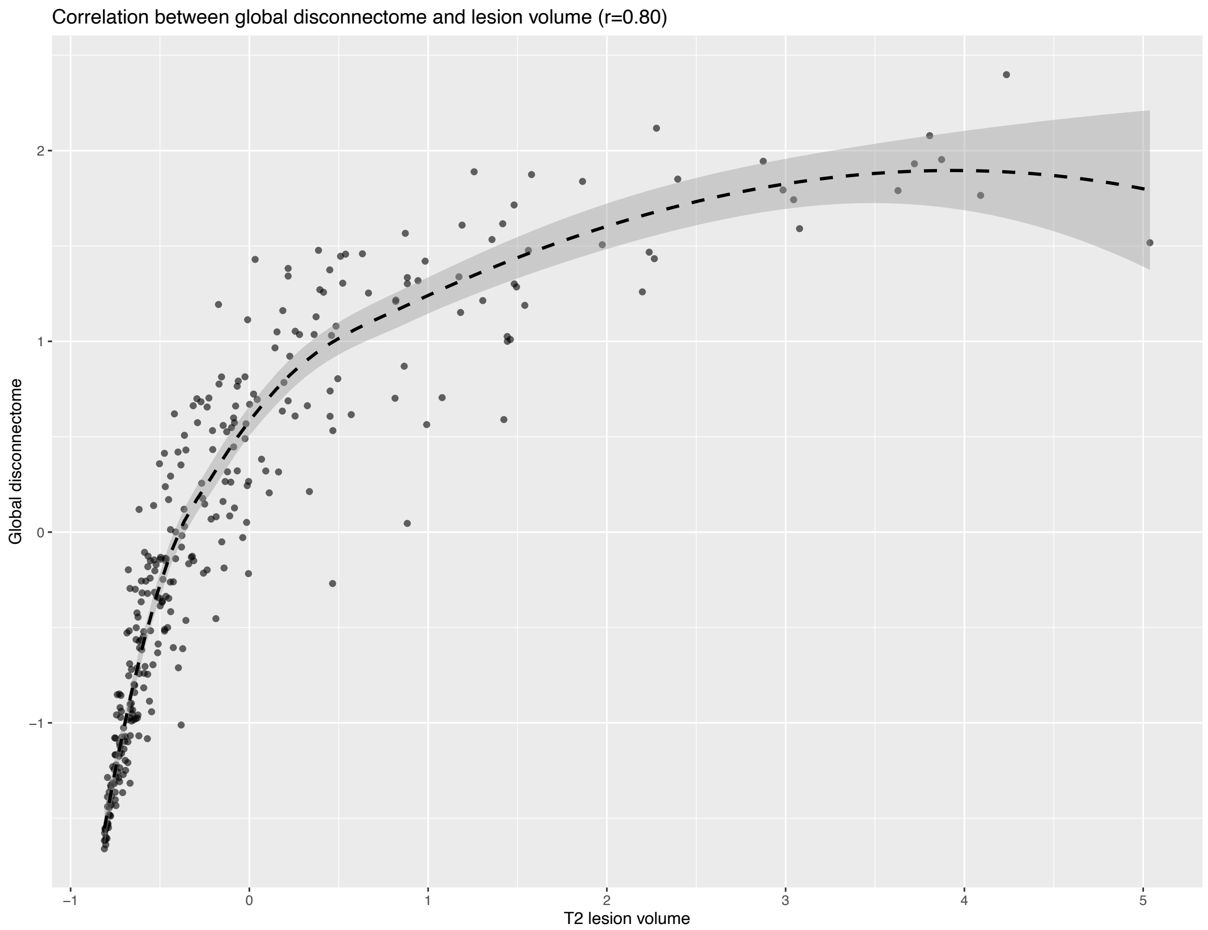


**Supplementary Figure 1 Visualization of the correlation between global disconnectome and T2 lesion volume**. Using normalized values, the correlation is high (r=0.80).

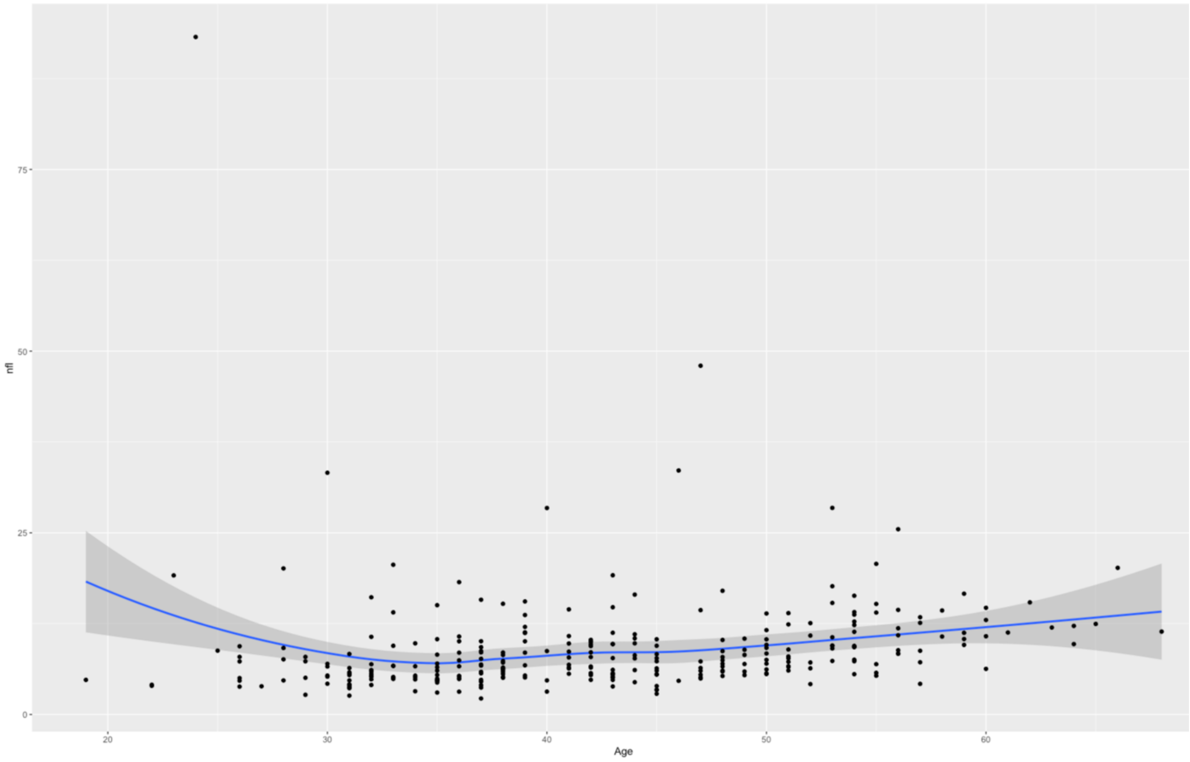


**Supplementary Figure 2 Scatter plot visualizing the distributions of sNfL levels across the complete sample with age on the x-axis.**

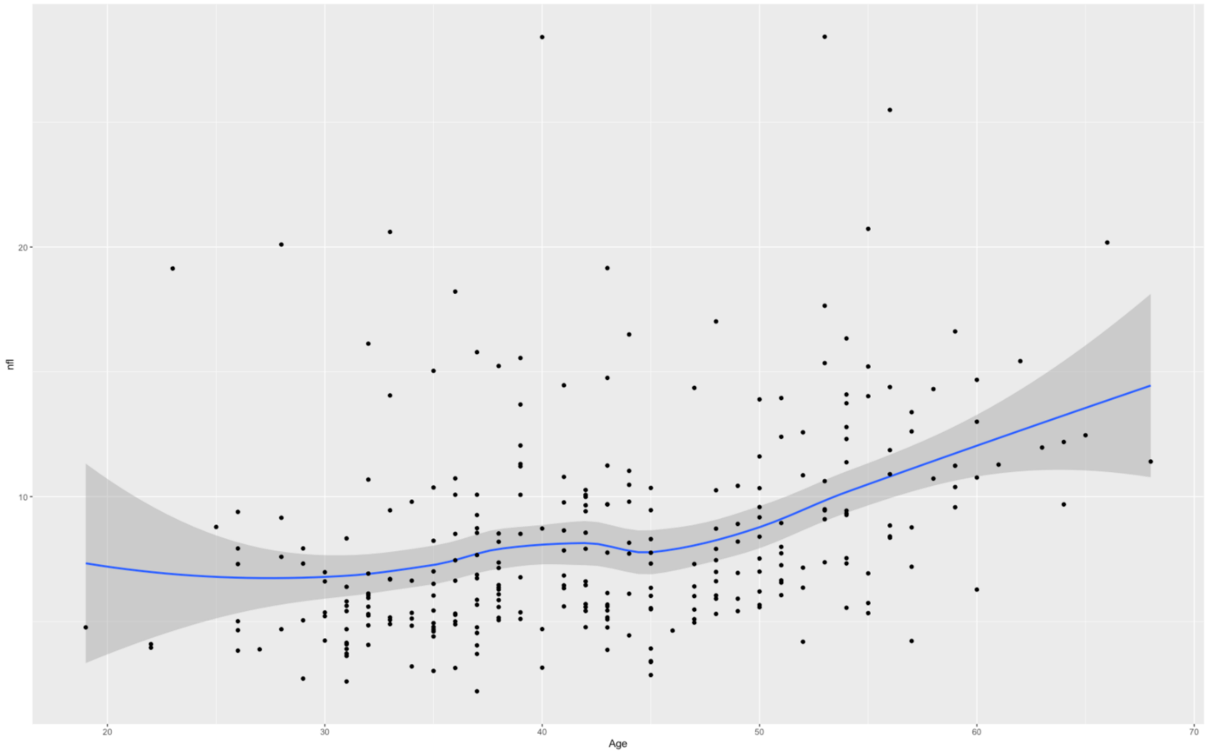


**Supplementary Figure 3 Scatter plot visualizing the distributions of sNfL levels across the sample with age on the x-axis, excluding outliers.**

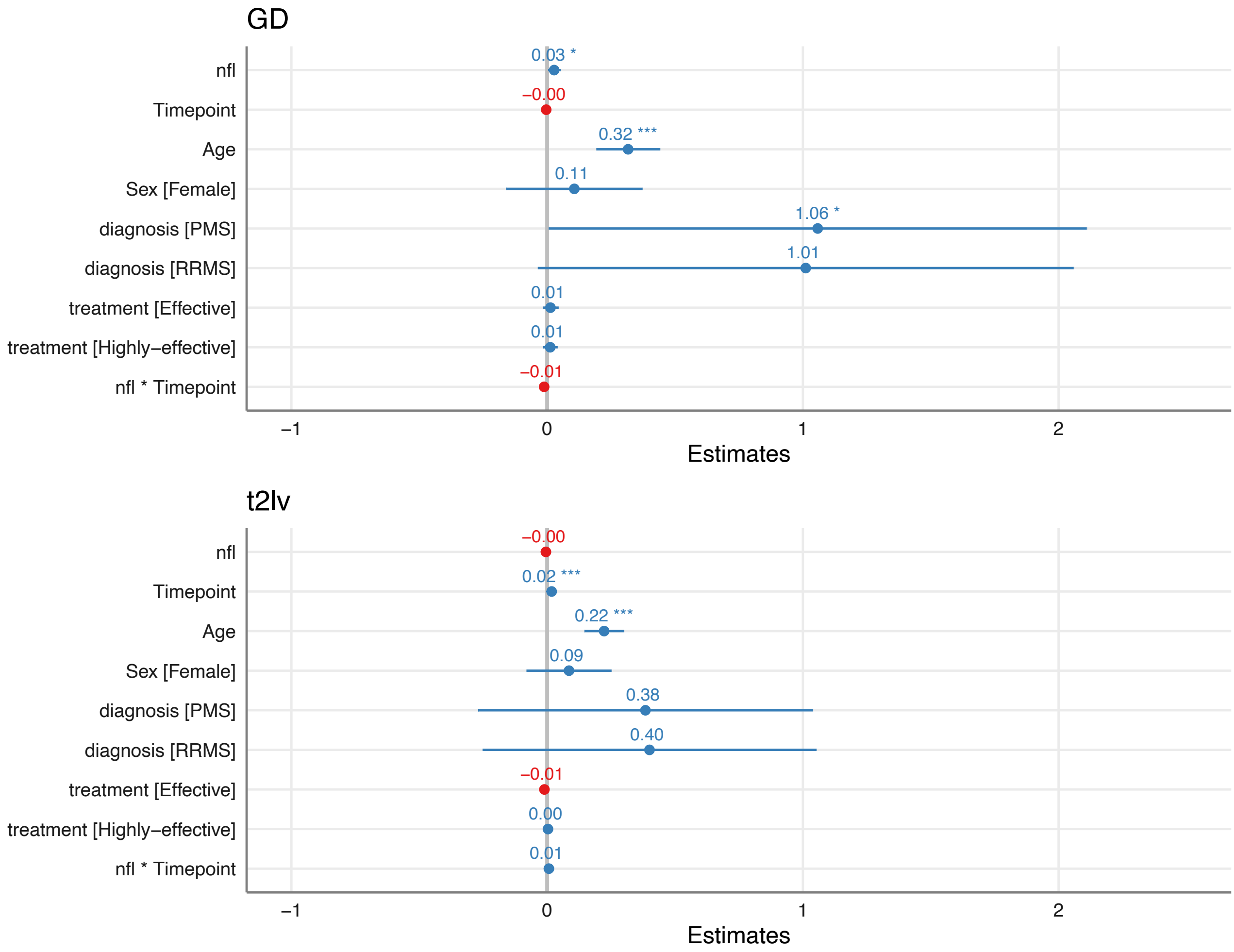


**Supplementary Figure 4 Visualization of fixed effects coefficients in linear mixed models. (A)** Higher GD was associated with higher NfL levels, higher age, and PMS diagnosis. **(B)** T2LV was found to increase over time and was associated with higher age. * $p < .05$, ** $p < .01$, *** $p < .001$.
